# AI-GUIDED ENDPOINT SELECTION FOR NEUROPROTECTION TRIALS IN GLAUCOMA

**DOI:** 10.1101/2025.08.23.25334286

**Authors:** Douglas R. da Costa, Rafael Scherer, Swarup Swaminathan, Henry Tseng, Felipe A. Medeiros

## Abstract

Standard Automated Perimetry (SAP) is the mainstay for monitoring glaucoma progression and has been accepted by the U.S. Food and Drug Administration (FDA) as a trial endpoint, but only under stringent criteria of ≥7 dB loss in five pre-specified test locations. Identifying such locations a priori has remained a major barrier for neuroprotection trials. We developed an attention-based graph neural network (GNN) to predict the visual field points most likely to deteriorate (High-5) using baseline SAP data. The model was trained in the Bascom Palmer Ophthalmic Registry (BPOR; 6,996 eyes, 5,405 patients, 40,914 tests) and externally validated in the Duke Glaucoma Registry (DGR; 5,211 eyes, 3,933 patients, 31,225 tests) and the University of Washington Humphrey Visual Field dataset (UWHVF; 2,030 eyes, 1,195 patients, 10,310 tests). In internal validation, the mean slope at High-5 points among progressors was −2.16±0.80 dB/year, compared to −0.55±0.44 dB/year for Low-5 and −1.02±0.40 dB/year for mean deviation (MD). Similar results were observed in DGR (–2.05 vs −0.45 vs −0.93 dB/year) and UWHVF (−2.32 vs −0.66 vs −1.14 dB/year). High-5 showed superior discrimination of progressors from non-progressors with areas under the ROC curve of 0.883, 0.898, and 0.937 across the three cohorts, consistently outperforming MD (0.871–0.911) and Low-5 (0.668–0.731). Nearly all progressing eyes exhibited a repeatable *≥*7 dB loss in average High-5 sensitivity during follow-up, compared to fewer than 30% when using MD. In sample size projections, High-5 increased the absolute effect size and lowered the *σ*^2^/Δ^2^ ratio, translating into an estimated 42% reduction in required trial size compared to MD. In conclusion, this GNN-based framework enables data-driven identification of high-risk SAP locations, aligning with regulatory definitions of progression while substantially improving trial efficiency and sensitivity to detect meaningful visual field change.

**Funding:** This work was supported in part by NIH R01 (EY036593) and by the Glaucoma Research Foundation (grant Endpoints2025MedeF).

## 1 Introduction

Glaucoma is a progressive optic neuropathy and a leading cause of irreversible blindness worldwide.^1^ Although lowering intraocular pressure (IOP) can slow disease progression, many patients deteriorate even with well-controlled IOP,^2,3^ emphasizing the need for alternative therapeutic strategies. Neuroprotective therapies that directly target retinal ganglion cell survival hold potential for preserving vision. However, their development has been hindered by the absence of reliable clinical trial endpoints to accurately assess disease progression.

Standard Automated Perimetry (SAP) remains the primary method for monitoring glaucoma progression, providing quantitative assessments of visual function over time. In addition to its established role in clinical care, SAP serves as a critical endpoint in clinical trials evaluating novel therapeutic strategies aimed at slowing or halting disease progression. The U.S. Food and Drug Administration (FDA) accepts SAP as a valid endpoint in Phase III clinical trials but requires a specific definition of progression: a 7 dB deterioration at a minimum of five pre-specified test locations.^4-6^ While this criterion enhances specificity and reduces variability, it presents practical challenges. Glaucomatous progression is often slow and spatially heterogeneous, making it difficult to satisfy this threshold using conventional, non-targeted approaches. Furthermore, there are currently no validated methods to prospectively identify which visual field locations are most likely to deteriorate, limiting the ability to optimize endpoint selection. Addressing these limitations through advanced analytical strategies is critical to enabling more efficient trials and accelerating the development of effective glaucoma therapies.

Recent advancements in artificial intelligence (AI), particularly deep learning, offer new opportunities to enhance the analysis of SAP data by efficiently handling its complexity. Graph Neural Networks (GNNs)^7^ provide a promising approach for analyzing visual field data by modeling test locations and their interrelationships as a graph structure. In GNNs, data points are represented as nodes connected by edges that reflect spatial or functional associations. This structure allows information to propagate across nodes, capturing the underlying patterns of glaucomatous damage that may be overlooked by simpler analytical techniques or conventional deep learning structures. The application of GNNs to SAP data enables a more comprehensive assessment that considers both individual test locations and their spatial dependencies, potentially improving diagnostic accuracy and predictive capability.

In this study, we developed and evaluated an attention-based GNN model to identify the visual field locations most likely to exhibit glaucomatous progression based on baseline SAP data. The attention mechanism within the GNN framework allows the model to focus on the most informative test locations, enhancing its ability to predict progression patterns. The proposed approach facilitates the identification of the five test points most likely to deteriorate, providing a data-driven method for pre-specifying endpoints in clinical trials. This framework aligns with stringent FDA requirements and has the potential to improve the efficiency and reliability of clinical trials for glaucoma therapies. Additionally, it offers pathways for optimizing current algorithms to detect visual field progression in clinical practice.

## 2 Methods

This study utilized de-identified electronic medical records and SAP test data from three sources: the Bascom Palmer Eye Institute (Bascom Palmer Ophthalmic Registry, BPOR), University of Miami, Florida;^8^ Duke University (Duke Glaucoma Registry, DGR), Durham, North Carolina;^9^ and the publicly available University of Washington Humphrey Visual Field (UWHVF) dataset, Seattle, Washington.^10^ The Institutional Review Boards at the University of Miami and Duke University approved the studies leading to the development of these registries and granted a waiver of informed consent due to their retrospective nature. The UWHVF dataset was de-identified in accordance with Institutional Review Board–approved protocols and Health Insurance Portability and Accountability Act (HIPAA) regulations. The study adhered to the tenets of the Declaration of Helsinki for research involving human subjects.

### 2.1 Participants and Eligibility Criteria

Participants were eligible if they had reliable SAP tests using the 24-2 strategy on the Humphrey Field Analyzer (HFA, Carl Zeiss Meditec Inc, Dublin, CA, USA), defined as fixation losses <33% and false positives <15%. Unreliable tests were excluded. Eyes were required to have at least four reliable SAP tests and a minimum follow-up period of one year. Glaucoma eyes had to show repeatable glaucomatous abnormality at baseline, defined by Glaucoma Hemifield Test (GHT) outside normal limits or pattern standard deviation (PSD) with P <5% on two consecutive tests. Baseline MD was required to be better than −15 dB to exclude eyes with advanced damage in which further progression with the 24-2 strategy would be difficult to detect. Follow-up was capped at 6 years, as predictions extending beyond this timeframe are unlikely to provide meaningful clinical guidance or inform endpoint selection in the context of prospective clinical trials. Eyes were excluded if they had ocular or systemic conditions that could affect visual field outcomes, including uveitis, retinal detachment, tumors, non-glaucomatous optic neuropathies, macular degeneration, maculopathy, amblyopia, or vascular occlusions.

The UWHVF dataset comprises both glaucoma and non-glaucoma cases, although most eyes with follow-up data are glaucomatous.^10^ Age, gender, and laterality were the only available clinical data. For consistency, the same inclusion criteria were applied to the UWHVF dataset, including the reliability criteria and requirement of at least four reliable SAP tests over a minimum of one year and a maximum of 6 years of follow-up.

### 2.2 Outcome Definition

Progression was defined as a clinically meaningful deterioration of *≥*7 dB in sensitivity in at least five test locations, relative to the average of the two baseline visual fields. Progression had to be confirmed by repeatable deterioration at the same locations on a subsequent test. The date of confirmation was recorded as the progression date. Notably, this criterion did not require pre-specification of the points where progression might occur. Instead, the goal of this study was to develop an algorithm capable of predicting such locations.

### 2.3 Model Development Cohort

Data from the BPOR were used for model development. After applying inclusion and exclusion criteria, 40,914 SAP tests from 6,996 eyes of 5,405 subjects with OAG were retained. The average follow-up was 4.9 ± 1.5 years, with 5.8 ± 1.9 tests per eye. Of these, 1,307 subjects (24.2%) showed confirmed progression in at least one eye and were included in model development. This focused sampling strategy was adopted to specifically train the model to learn the spatial patterns of deterioration in eyes that had actually progressed. By excluding non-progressing eyes from the training phase, we ensured that the model focused on predicting the locations most likely to deteriorate in progressing eyes. The cohort was split at the patient level into 70% for training and 30% for internal validation. **Table 1** summarizes patient and eye-level characteristics.

**Table 1:** Demographic and clinical characteristics of participants in the internal and external validation cohorts.

| Characteristic | BPOR | DGR | UWHVF |
| --- | --- | --- | --- |
| Number of patients, n | 5405 | 3933 | 1195 |
| Number of eyes, n | 6996 | 5211 | 2030 |
| Total number of tests, n | 40914 | 31225 | 10310 |
| Age, years | 66.4 (12.8) | 64.2 (14.1) | 63.1 (14.0) |
|  | 67.8 (59.7–75.2) | 66.5 (57.4–73.7) | 64.9 (56.3–72.6) |
| Female sex, n (%) | 3204 (59.3%) | 2281 (58.0%) | 545 (45.6%) |
| Race: Asian, n (%) | 83 (1.5%) | 158 (4.0%) | N/A |
| Race: Black, n (%) | 1021 (18.9%) | 1008 (25.6%) | N/A |
| Race: White, n (%) | 4301 (79.6%) | 2767 (70.4%) | N/A |
| Hispanic ethnicity, n (%) | 1867 (34.5%) | 71 (1.8%) | N/A |
| Mean Deviation, dB | -5.6 (3.7) | -4.9 (3.7) | -4.6 (3.2) |
|  | -4.8 (-7.9– -2.8) | -3.9 (-6.8– -2.1) | -3.8 (-5.9– -2.4) |
| Pattern Standard Deviation, dB | 5.5 (3.4) | 5.0 (3.4) | 3.3 (2.5) |
|  | 4.2 (2.7–7.6) | 3.6 (2.3–7.1) | 2.2 (1.6–4.1) |
| Tests per eye, n | 5.8 (1.9) | 6.0 (2.2) | 5.1 (1.1) |
|  | 5.0 (4.0–7.0) | 5.0 (4.0–7.0) | 5.0 (4.0–6.0) |
| Follow-up duration, years | 4.9 (1.5) | 4.7 (1.5) | 4.5 (1.4) |
|  | 5.1 (3.7–6.1) | 4.9 (3.5–6.0) | 4.6 (3.4–5.6) |
| Eyes with progression, n (%) | 1453 (20.8%) | 903 (17.3%) | 171 (8.4%) |

Data are mean (SD), n (%), or median (IQR), as indicated. For continuous variables, both mean (SD) and median (IQR) are reported in separate rows when available. BPOR = Bascom Palmer Ophthalmic Registry. DGR = Duke Glaucoma Registry. UWHVF = University of Washington Humphrey Visual Field dataset. N/A = not available.

### 2.4 Graph Neural Network Architecture

We developed a GNN architecture to analyze baseline visual fields and identify locations most likely to progress. Each SAP test location was modeled as a node, with edges representing anatomically-informed spatial relationships. The graph consisted of 52 nodes (standard test points), excluding the two above and below the blind spot. Edges were restricted to points within the same hemifield and limited by horizontal and vertical distance thresholds to preserve local connectivity along arcuate pathways. This resulted in a sparse graph structure aligned with known patterns of retinal nerve fiber organization and glaucomatous damage.

For each progression event in our dataset, we prepared input features using the average of the total deviation (TD) values of the first two visual field tests (baseline tests) for each location. This baseline averaging helps reduce measurement variability and provides a more stable starting point for progression prediction.

The model architecture was based on a Graph Attention Network (GAT)^11^ with multiple attention heads per layer to capture spatial relationships among test locations. It used a hierarchical structure, starting with 128 hidden dimensions and progressively reducing feature size. Batch normalization, decreasing dropout, and skip connections were incorporated to stabilize training and preserve low-level features. A global attention readout layer allowed each node to integrate information from all others, capturing long-range dependencies. Its output was combined with the original features through a residual connection and passed through a final linear layer for prediction. The model was trained using a ranking loss function, encouraging accurate prioritization of the locations most likely to exhibit future progression. Training used the AdamW optimizer with cosine annealing learning rate scheduling and early stopping based on validation rank performance.

### 2.5 Point Selection Strategy

To identify the most likely locations to progress, we implemented a point selection strategy combining model-derived probabilities, uncertainty estimation, and anatomical context. For each test, multiple stochastic forward passes using dropout were performed to estimate mean progression probabilities and predictive uncertainty at each location. Points were then scored using a weighted combination of probability, certainty, and neighborhood features.

The five locations with the highest composite scores were selected as High-5, reflecting both statistical likelihood and anatomical plausibility. The five lowest-scoring locations (Low-5) were selected for comparison.

### 2.6 External Validation

The trained model was applied to the DGR and UWHVF datasets without retraining. For each eye, High-5 and Low-5 were identified by applying the model to the average TD from two baseline tests. The average TD for each metric was tracked across all follow-up visits. Linear mixed models^12^ were used separately for each dataset to estimate rates of change (in dB/year) for High-5, Low-5, and global MD.

### 2.7 Statistical Analysis

To assess the spatial performance of the GNN pointwise predictions, we used two complementary metrics. First, we calculated the mean rank of progressing locations, defined as the average position assigned to these points when all 52 visual field locations were ordered by the model’s predicted probability of progression. Lower values indicate better performance. We defined strong performance as an average rank below 15. This threshold reflects a scenario in which the majority of the progressing points per eye are ranked within the top quartile (the top 13 of 52 locations) and reflects a meaningful improvement over the random expectation of 26.5. Second, we computed a neighborhood-based hit rate, defined as the proportion of eyes in which at least one progressing point, or one of its immediate spatial neighbors, appeared among the top five ranked locations.

To assess the discriminative ability of the High-5 locations, we constructed receiver operating characteristic (ROC) curves to differentiate progressors from non-progressors. The area under the ROC curve (AUC) was used as a measure of overall classification performance, where a value of 1.0 indicates perfect discrimination and 0.5 corresponds to chance-level performance. Confidence intervals were estimated using non-parametric bootstrapping with 10,000 resamples, performed at the subject level to account for correlation between fellow eyes.

We also performed a time-to-event analysis to assess the cumulative probability of developing a significant change in High-5 in eyes that eventually developed progression. For this analysis, an event was defined as a repeatable (two consecutive tests) decline of *≥*7 dB in the mean sensitivity of the High-5 points, from baseline. This threshold was selected based on FDA-accepted criteria used in prior clinical trials involving perimetric endpoints. A similar definition was applied to MD and Low-5 to enable direct comparison. Kaplan-Meier survival curves were generated for each group, and eyes not reaching the event threshold were censored at their last follow-up. Survival distributions were compared using the log-rank test.

Model training was conducted using PyTorch. Statistical analyses were performed in Stata (StataCorp, College Station, TX). A two-sided alpha of 0.05 was used to determine statistical significance.

## 3 Results

A total of 82,449 SAP tests from 14,237 eyes of 10,533 patients were included across three datasets (**Table 1**). The BPOR dataset comprised 6,996 eyes from 5,405 patients, with a mean age of 66.4 ± 12.8 years, baseline MD of −5.6 ± 3.7 dB, and follow-up of 4.9 ± 1.5 years. The DGR dataset included 5,211 eyes from 3,933 patients, with a mean age of 64.2 ± 14.1 years, MD of −4.9 ± 3.7 dB, and follow-up of 4.7 ± 1.5 years. The UWHVF dataset included 2,030 eyes from 1,195 patients, with a mean age of 63.1 ± 14.0 years, MD of −4.6 ± 3.2 dB, and follow-up of 4.5 ± 1.4 years. The proportion of progressing eyes was 20.8% in BPOR, 17.3% in DGR, and 8.4% in UWHVF.

The GNN model demonstrated strong ability to predict progression locations. In the internal validation set, the mean rank of progressing points was 14.0 (95% CI: 13.5–14.5), and in the DGR external test set, 13.9 (95% CI: 13.5–14.5) and 15.7 (95% CI: 14.4 – 17.2) in the UWHVF, all significantly better than expected under random ranking (26.5; p < 0.001 for all comparisons). Using a neighborhood-based eye-level hit rate metric, the Top 5 performance was 88.3% (95% CI: 85.9–91.6%) in the internal validation set and 89.7% (95% CI: 87.6–91.6%) in the external test set DGR and 90.1% (95% CI: 85.4-94.2%) in external set UWHVF.

Among eyes that progressed during follow-up in the internal validation set, the mean TD slope at High-5 locations was -2.16 ± 0.80 dB/year, significantly steeper than at Low-5 (-0.55 ± 0.44 dB/year; P < 0.001) and global MD (-1.02 ± 0.40dB/year; P < 0.001). The distribution of slopes across selection strategies is shown in **Figure 1**, and corresponding summary statistics are provided in **Table 2**.

**Table 2:** Rates of Visual Field Change and Discriminative Performance of High-5, Low-5, and MD Metrics in Progressor and Non-progressor Eyes.

| Metric | Internal Validation (BPOR) |  | External Dataset 1 (DGR) |  | External Dataset 2 (UWHVF) |  |
| --- | --- | --- | --- | --- | --- | --- |
|  | Progressors | Non-progressors | Progressors | Non-progressors | Progressors | Non-progressors |
| Number of patients, n | 423 | 1336 | 823 | 3394 | 160 | 1136 |
| Number of eyes, n | 475 | 1613 | 903 | 4308 | 171 | 1859 |
| **High-5 points slope** |  |  |  |  |  |  |
| Mean (SD), dB/year | -2.16 (0.80) | -0.22 (0.52) | -2.05 (0.82) | -0.16 (0.47) | -2.32 (1.04) | -0.16 (0.30) |
| Median (Q1–Q3), dB/year | -2.17 (-2.72 – -1.70) | -0.12 (-0.51–0.14) | -2.08 (-2.62 – -1.50) | -0.07 (-0.42–0.18) | -2.31 (-2.99 – -1.65) | -0.08 (-0.31–0.06) |
| Slopes < -1 dB/year, n (%) | 434 (91.4%) | 129 (8.0%) | 806 (89.3%) | 256 (5.9%) | 155 (90.6%) | 37 (2.0%) |
| AUC (95% CI) | 0.883 (0.865–0.900) |  | 0.898 (0.888–0.909) |  | 0.937 (0.917–0.957) |  |
| **Low-5 points slope** |  |  |  |  |  |  |
| Mean (SD), dB/year | -0.55 (0.44) | -0.20 (0.42) | -0.45 (0.50) | -0.10 (0.30) | -0.66 (0.89) | -0.12 (0.16) |
| Median (Q1–Q3), dB/year | -0.45 (-0.75 – -0.23) | -0.12 (-0.30–0.05) | -0.35 (-0.68 – -0.13) | -0.06 (-0.20–0.07) | -0.42 (-0.94 – -0.13) | -0.10 (-0.18 – -0.02) |
| Slopes < -1 dB/year, n (%) | 71 (14.9%) | 69 (4.3%) | 111 (12.3%) | 59 (1.4%) | 37 (21.6%) | 5 (0.3%) |
| AUC (95% CI) | 0.677 (0.649–0.702) |  | 0.668 (0.649–0.687) |  | 0.731 (0.688–0.773) |  |
| **Mean Deviation (MD) slope** |  |  |  |  |  |  |
| Mean (SD), dB/year | -1.02 (0.40) | -0.18 (0.43) | -0.93 (0.44) | -0.10 (0.22) | -1.14 (0.72) | -0.11 (0.17) |
| Median (Q1–Q3), dB/year | -1.01 (-1.25 – -0.75) | -0.12 (-0.32–0.06) | -0.91 (-1.15 – -0.65) | -0.06 (-0.20–0.04) | -1.07 (-1.45 – -0.73) | -0.09 (-0.19 – -0.01) |
| Slopes < -1 dB/year, n (%) | 245 (51.6%) | 59 (3.7%) | 354 (39.2%) | 24 (0.6%) | 94 (55.0%) | 3 (0.2%) |
| AUC (95% CI) | 0.871 (0.854–0.888) |  | 0.875 (0.864–0.887) |  | 0.911 (0.886–0.933) |  |

Data are mean (SD), median (IQR), or n (%), as indicated. Area under the curve (AUC) values reflect discrimination between progressors and non-progressors based on slope from linear mixed-effects models. BPOR = Bascom Palmer Ophthalmic Registry. DGR = Duke Glaucoma Registry. UWHVF = University of Washington Humphrey Visual Field dataset. High-5 = five locations with highest predicted risk; Low-5 = five lowest-risk locations; MD = mean deviation.

**Figure 1.**
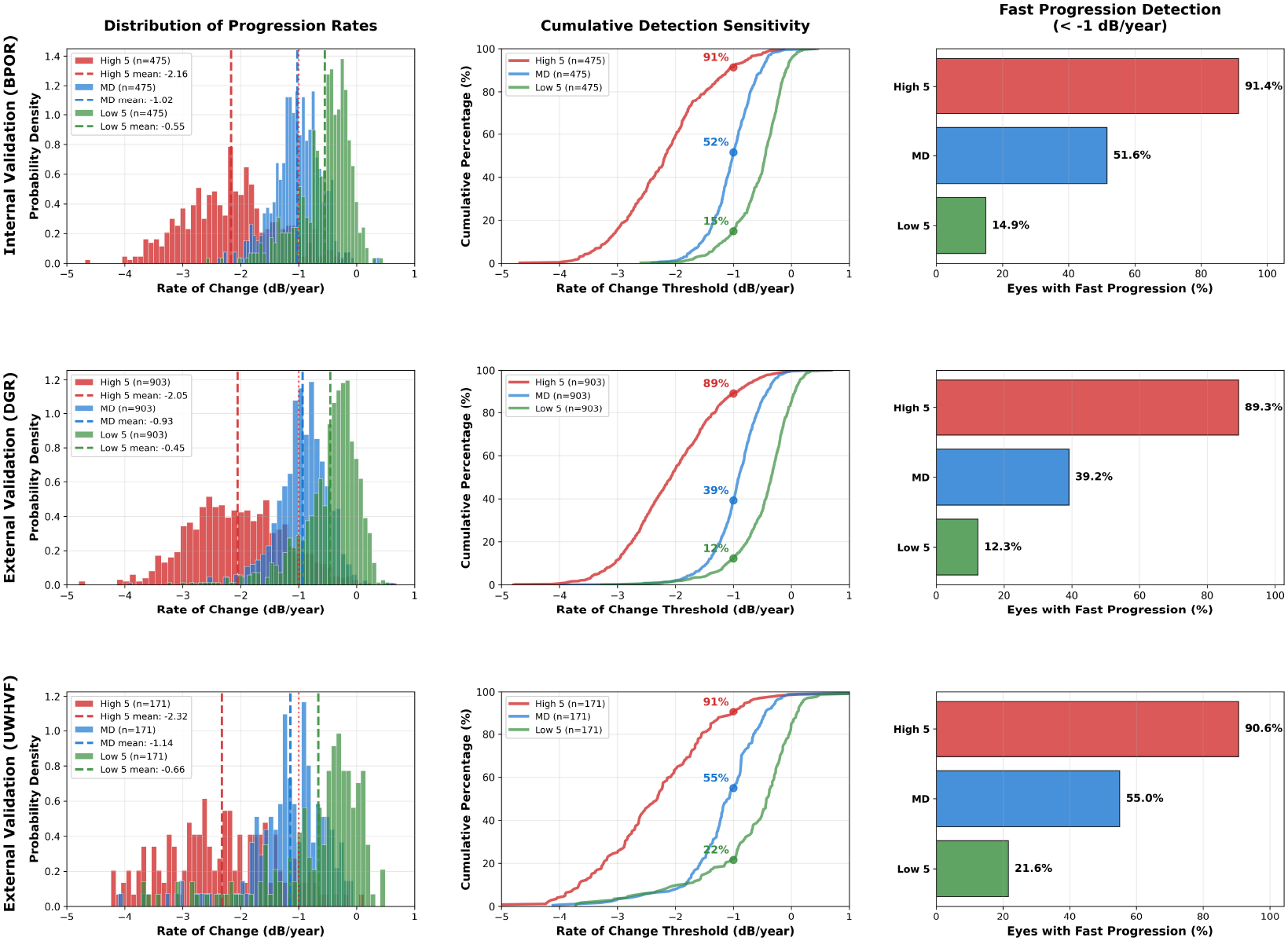
Distribution of progression rates and detection of fast progressors across internal and external validation cohorts. Histograms (left) show the distribution of rates of change at High-5, MD, and Low-5 locations in progressing eyes from the internal validation (BPOR) and external validation cohorts (DGR and UWHVF). Middle panels display cumulative detection curves showing the proportion of eyes exceeding a given progression threshold for each metric. Right panels show the proportion of progressors with rapid progression (< −1 dB/year) under each metric. High-5 slopes were 4–5 times faster than Low-5 and approximately twice as fast as MD. High-5 consistently captured a greater proportion of rapid progressors across all datasets.

In the DGR external validation cohort, mean slopes were similarly steeper at High-5 locations among progressors, with an average of −2.05 ± 0.82 dB/year, compared to −0.45 ± 0.50 for Low-5 (P < 0.001) and −0.93 ± 0.44 for MD (P < 0.001). Slope distributions are shown in **Figure 1**, with High-5 slopes exhibiting a pronounced leftward skew, reflecting that a greater proportion of eyes experienced rapid functional decline at the locations identified as high probability by the model, as compared to Low-5 and MD. In fact, the proportion of progressors with High-5 slopes faster than −1 dB/year was 89.3%, compared to 12.3% and 39.2% for Low-5 and MD, respectively (**Table 2**).

Similar results were seen for the UWHVF external validation cohort 2 (**Table 2** and **Figure 1**). Mean High-5 slopes (-2.32 ± 1.04 dB/year) were much steeper than Low-5 (-0.66 ± 0.89 dB/year; P<0.001) and MD (-1.14 ± 0.72 dB/year; P<0.001). Also, the proportion of progressors with High 5 slopes faster than -1dB/year was 90.6% for High-5 compared to only 21.6% for Low-5 and 55% for MD.

**ROC analysis** for discriminating progressors from non-progressors yielded very similar results for the internal and both external validation cohorts (**Figure 2**). In the external validation cohort DGR, the AUC was 0.898 (95% CI: 0.888 – 0.909) for High-5, which was statistically significantly higher than Low-5 (0.668, 95% CI: 0.649 – 0.687; P < 0.001) and MD (0.875, 95% CI: 0.864 – 0.887; P < 0.001). In the external validation cohort UWHVF, the AUC for High-5 (0.937; 95% CI: 0.917 – 0.957) was also significantly better than the one for Low-5 (0.731; 95% CI: 0.688 – 0.773; P<0.001) and MD (0.911; 95% CI: 0.886 – 0.933; P=0.003).

**Figure 2.**
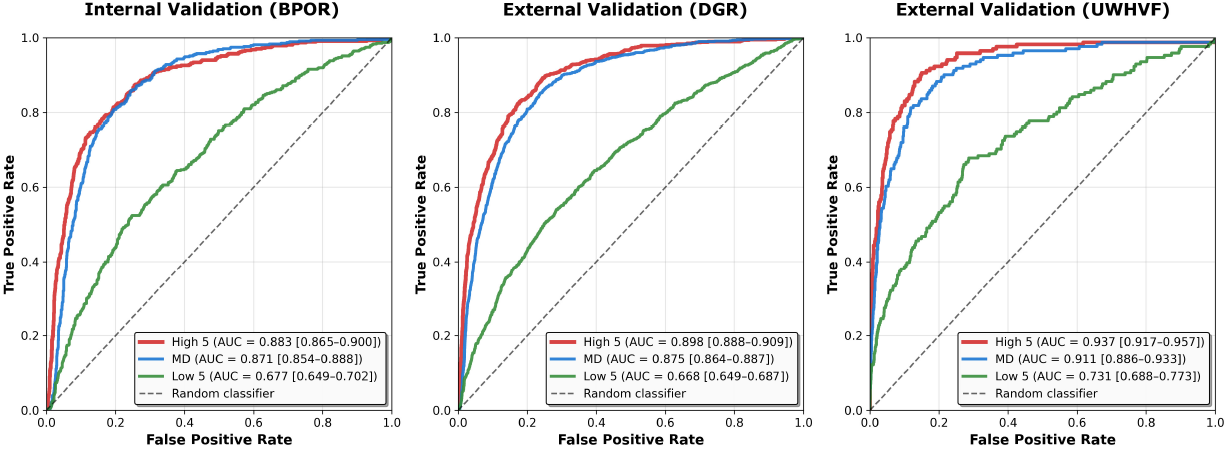
Receiver operating characteristic curves for discrimination of progressors versus non-progressors. ROC curves comparing High-5, MD, and Low-5 metrics across internal (BPOR) and external (DGR, UWHVF) validation cohorts. High-5 showed superior discriminative performance across all datasets, with AUCs ranging from 0.883 to 0.937, consistently outperforming both MD and Low-5 (p < 0.001 for all comparisons). AUC values and 95% confidence intervals are shown in each panel.

Figure 3. shows scatterplots of High-5 versus MD slopes for each one of the validation datasets, along with the line of equality. Inspection of the plots shows that almost all points are located below the line of equality, that is, with faster rates of High-5 compared to those of MD in the same eye.

**Figure 3.**
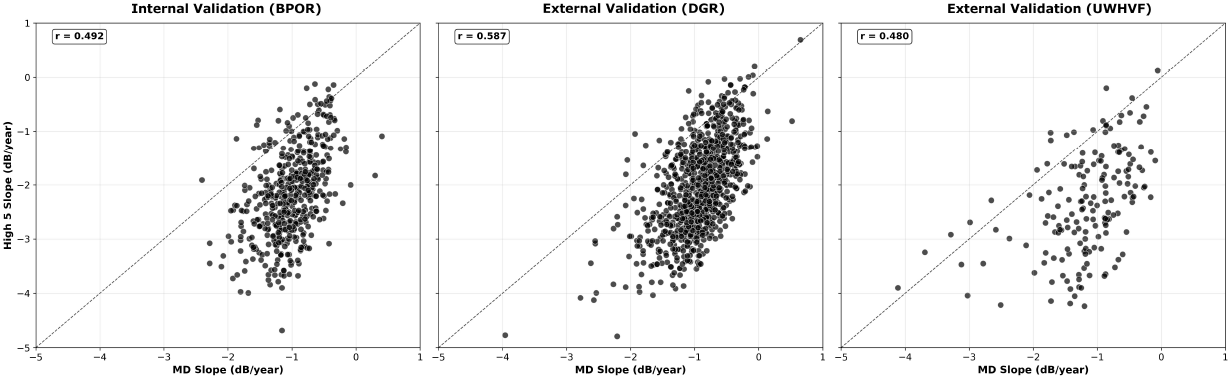
Distribution of progression rates and detection of fast progressors across internal and external validation cohorts. Histograms (left) show the distribution of rates of change at High-5, MD, and Low-5 locations in progressing eyes from the internal validation (BPOR) and external validation cohorts (DGR and UWHVF). Middle panels display cumulative detection curves showing the proportion of eyes exceeding a given progression threshold for each metric. Right panels show the proportion of progressors with rapid progression (< −1 dB/year) under each metric. High-5 slopes were 4–5 times faster than Low-5 and approximately twice as fast as MD. High-5 consistently captured a greater proportion of rapid progressors across all datasets.

Figure 4. shows Kaplan-Meier curves illustrating the cumulative probability of developing a repeatable decline of 7dB in the average sensitivity of the High-5 points, as compared to the other point selection strategies. In all cohorts, eyes that eventually progressed almost always developed the 7dB repeatable decline in the average sensitivity of High-5 points, with cumulative probabilities of 95%, 94% and 100% for the internal validation, external cohort DGR and external cohort UWHVF. In contrast, cumulative probabilities were much lower for MD and Low-5 metrics.

**Figure 4.**
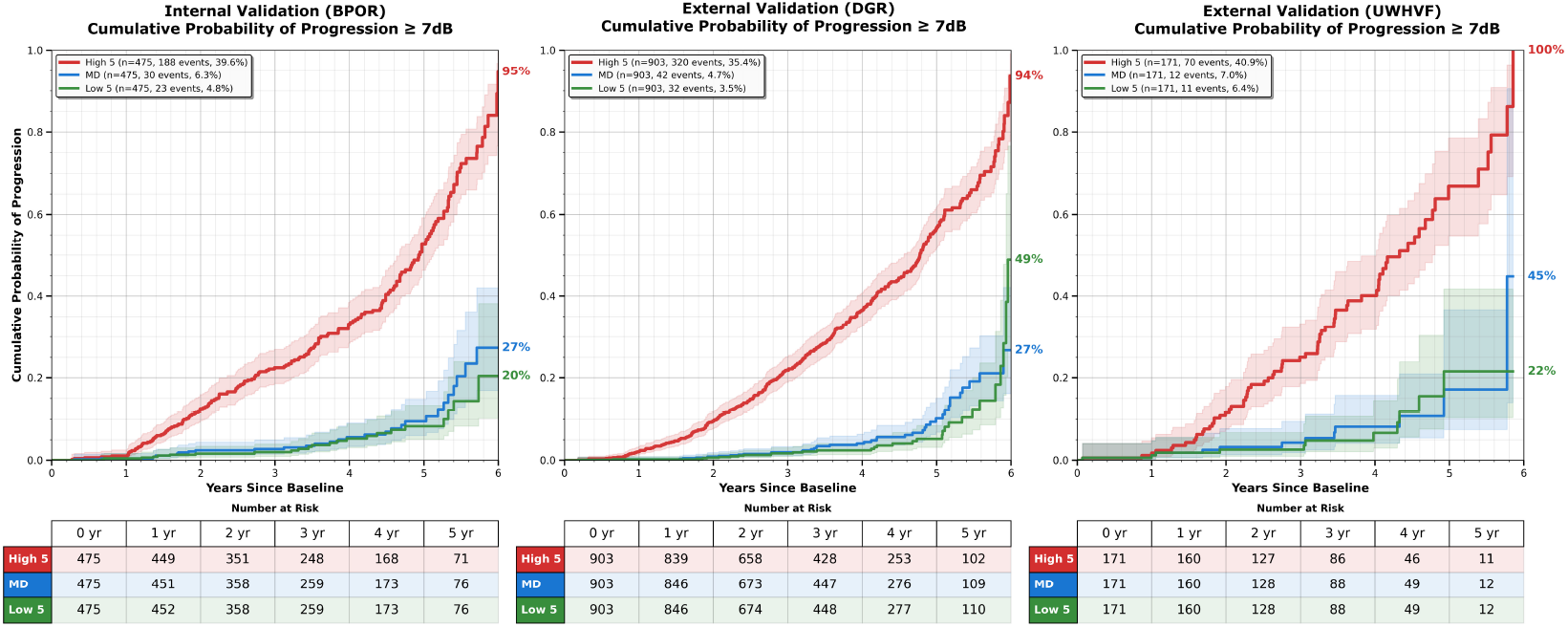
Time-to-event analysis of repeatable *≥*7 dB decline. Kaplan–Meier curves showing cumulative probability of developing a repeatable *≥*7 dB sensitivity loss over time at High-5, MD, and Low-5 locations in progressing eyes across validation cohorts. High-5 showed markedly higher event rates of 95%, 94%, and 100% in BPOR, DGR, and UWHVF, respectively, compared to MD and Low-5. Shaded regions indicate 95% confidence intervals. Numbers at risk are presented below each timepoint.

Figure 5. presents the spatial distribution of High-5 selection frequency across the visual field in the validation cohorts. Larger circles correspond to points that were selected more often as part of High-5. Highly selected points were generally in the inferior and superior arcuate regions and nasal area, as expected. The color code indicates the rate of change, showing that the selected points were indeed those that generally progressed faster.

**Figure 5.**
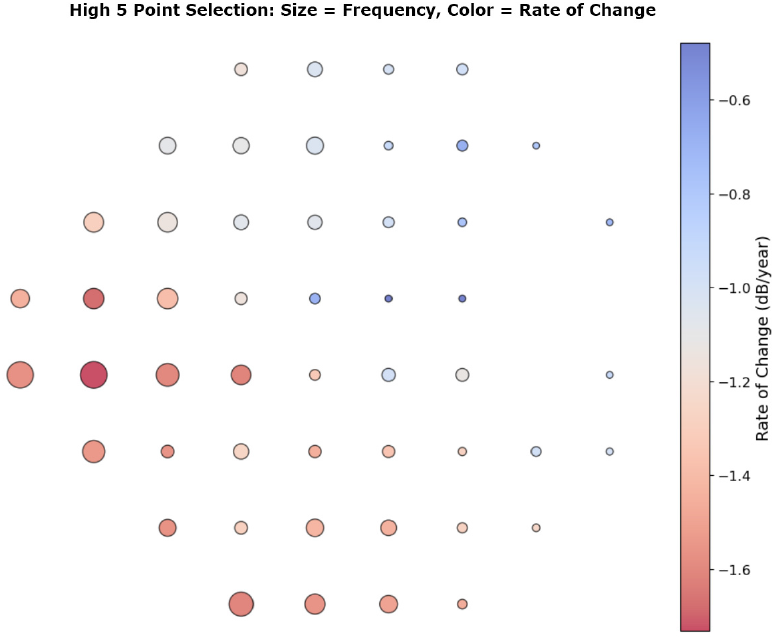
Spatial distribution of High-5 selection frequency and rate of change. Each circle represents a visual field location. Circle size indicates the frequency with which the point was selected as part of High-5 across progressing eyes in validation datasets. Color represents the average rate of change at each location, with warmer tones indicating faster deterioration. Most frequently selected points were concentrated in the superior and inferior arcuate and nasal regions.

Figure 6. shows representative examples of progressing eyes from the external validation cohorts, each with distinct baseline visual field patterns. For each eye, the left panel displays the two baseline visual fields and the two follow-up tests confirming progression. The right panel presents a heatmap of the model-predicted probability of progression at each location, with the five most likely points to progress (High-5) indicated by red circles. These examples illustrate the model’s ability to adapt to varying damage patterns and highlight specific high-risk regions within the visual field.

**Figure 6.**
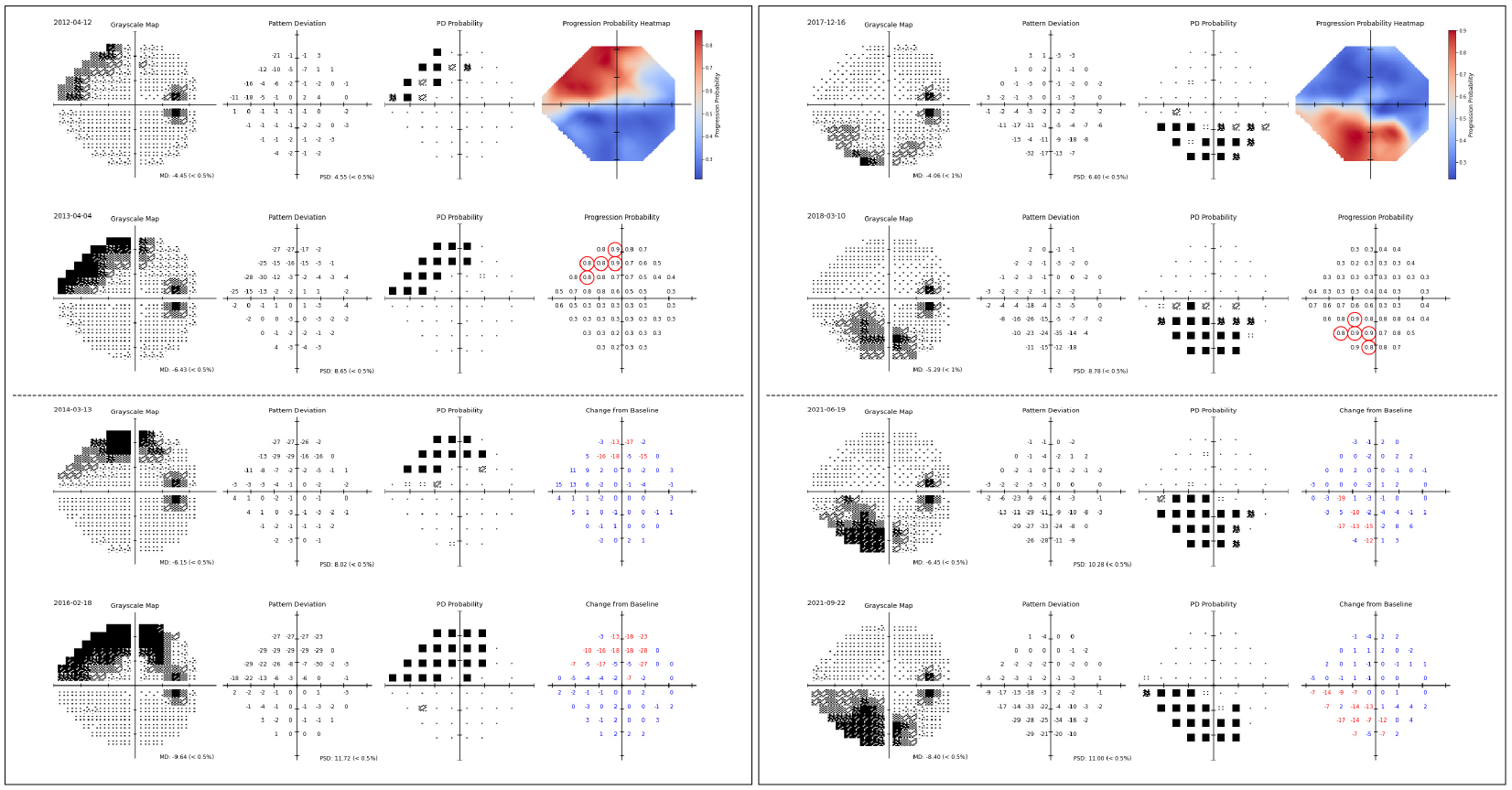
Representative examples of progression prediction in individual eyes. Two progressing eyes from the external validation cohorts are shown. For each eye, the left column displays two baseline and two follow-up SAP grayscale maps, pattern deviation plots, and probability maps. The right column shows heatmaps of the model-predicted progression probability at each location and the five most likely locations to progress (High-5, circled in red). High-5 selections aligned with regions of confirmed deterioration on follow-up testing, illustrating the model’s adaptability to diverse patterns of glaucomatous damage.

## 4 Discussion

In this study, we developed and validated an attention-based graph neural network model to predict which visual field locations are most likely to exhibit glaucomatous progression based solely on baseline perimetry data. The model showed consistent accuracy across internal and external validation cohorts, with AI-selected high-risk points displaying substantially faster decline than low-risk locations and traditional global measures such as MD. These results suggest that machine learning can uncover individualized patterns of vulnerability, offering a pathway to improve both clinical monitoring and the design of more efficient neuroprotection trials.

Across the internal and external validation datasets, average slopes of change at the High-5 locations were 4 to 5 times steeper than those at the Low-5 locations in eyes that showed progression (–2.05 to −2.32 dB/year vs. −0.45 to −0.66 dB/year). Critically, among non-progressing eyes, slopes were similar across High-5 and Low-5, indicating that the model does not merely select pre-existing damaged points but rather identifies regions at highest risk of future deterioration. The distinction between High-5 and Low-5 was further supported by ROC analysis, with area under the curve (AUC) values ranging from 0.883 to 0.937 for High-5, compared to only 0.668 to 0.731 for Low-5 (p < 0.001). This marked difference in discriminative performance demonstrates the model’s ability to effectively rank progression risk across visual field locations, with clear separation between high-risk and low-risk regions.

Notably, High-5 slopes were also approximately twice as fast, on average, as those of MD. As illustrated in Figure 1, nearly all progressing eyes exhibited faster rates of decline at High-5 locations compared to MD, often by a large amount. Although MD has traditionally been used to assess rates of visual field change in glaucoma, it has several important limitations. As a global index, MD is sensitive to diffuse changes caused by non-glaucomatous factors such as cataracts or ocular surface disease, which can artificially exaggerate or mask true progression. To improve detection of localized change, pointwise event-based strategies such as the Guided Progression Analysis (GPA) have been developed. However, GPA evaluates each visual field test location independently and treats all locations equally, without incorporating prior knowledge of anatomical susceptibility. This agnostic approach limits efficiency by including regions that are unlikely to progress, thereby increasing noise. In fact, when matched for specificity, GPA does appear to show increased sensitivity for progression compared to a global metric such as MD.^13^ Prior studies have shown that glaucomatous progression typically occurs through deepening or expansion of existing scotomas,^14-16^ yet formalizing this knowledge into predictive algorithms has been challenging. Our model addresses this limitation through the use of a graph-based deep learning framework that explicitly captures the spatial relationships between visual field locations. By learning patterns of glaucomatous damage and their association with future progression, the model was able to identify the points most likely to deteriorate, enhancing sensitivity to true progression while reducing the confounding of less relevant locations.

Our results demonstrated that the High-5 approach significantly outperformed MD in discriminating progressors from non-progressors across the 2 external validation cohorts. In the DGR and UWHVF datasets, the areas under the ROC curves for High-5 were 0.898 and 0.937, respectively, compared to 0.875 and 0.911 for MD. Although the absolute difference is not large, this is a remarkable result given that High-5 relies on only five pre-specified points, whereas MD summarizes information from the entire visual field. These findings suggest the potential for developing targeted perimetric strategies that focus more intensively on the most vulnerable locations, rather than testing all 52 points at every visit. Such an approach could substantially reduce testing time, improve patient comfort and adherence, and lower costs. Targeted home-based perimetry may also become feasible, with full 24-2 testing reserved for less frequent in-office assessments. While promising, these strategies will require rigorous validation to ensure they maintain diagnostic sensitivity and reliability across a broad spectrum of disease severity.

A key motivation for developing the targeted High-5 approach was to address a major limitation in the field of glaucoma, which is the lack of sensitive and specific functional endpoints suitable for neuroprotection clinical trials. The only large-scale neuroprotection trial in glaucoma to date, the memantine trial conducted in the early 2000s, enrolled over 2,000 patients followed for four years, yet failed to demonstrate a treatment benefit.^17^ One likely contributing factor was the use of the event-based Glaucoma Change Probability (GCP) endpoint, an earlier precursor of the GPA algorithm, which is insensitive and statistically inefficient as an endpoint. Recent studies have shown that the use of trend-based analyses, such as rates of change in MD, instead of event-based algorithms, could dramatically reduce sample size requirements for clinical trials.^18-20^ Wu and Medeiros^18^ demonstrated that a two-year trial targeting a 30% slowing in MD slope would require approximately 276 eyes per arm, an 85–90% reduction relative to traditional event-based designs. However, as previously noted, MD is a global metric and prone to noise from non-glaucomatous factors. This may be especially problematic in neuroprotection trials, where investigational treatments, including intravitreal therapies, may induce lens changes or other confounders. As a result, the FDA has not formally endorsed MD slope as a standalone functional endpoint, citing concerns about its specificity to glaucomatous damage. The High-5 approach addresses this limitation by targeting high-risk locations based on individualized patterns of glaucomatous damage, potentially offering a more sensitive and efficient functional endpoint for clinical trials.

Analysis of progression rates in the external validation cohorts suggests that using the High-5 metric as a functional endpoint could substantially improve the statistical efficiency of clinical trials. Among eyes that progressed, the average rate of change at High-5 locations across both external datasets was −2.19 dB/year, compared to −1.04 dB/year for MD. For a treatment aiming to slow progression by 30%, this corresponds to an absolute effect size (Δ) of 0.66dB/year with High-5 and 0.31 dB/year with MD, representing more than twice the magnitude of change. Although variability in High-5 slopes was greater, as expected due to averaging only a few points, the gain in effect size outweighed this, resulting in a lower *σ*^2^*/*Δ^2^ ratio: 2.00 for High-5 versus 3.46 for MD. As sample size is directly proportional to this ratio^21^, these values imply a 42% reduction in required sample size when using High-5 instead of MD to detect the same relative treatment effect. While these estimates are based on progressor-only data and assume population homogeneity, they highlight the potential of High-5 to improve trial efficiency. Future simulations incorporating real-world parameters such as testing frequency, dropout, and enrichment strategies will be needed to refine these projections.

In addition to continuous slope-based endpoints, it is important to consider binary, event-based criteria that align with regulatory precedents. The FDA has previously accepted a *≥*7 dB sensitivity change in five or more pre-specified test locations as a primary functional endpoint in gene therapy trials for RPGR-associated retinitis pigmentosa, including the AGTC-501^5^ and cotoretigene toliparvovec^22,23^ studies. A similar framework could be applied in glaucoma by evaluating average sensitivity across the five high-risk locations (High-5) identified at baseline. In our analysis, nearly all progressing eyes exhibited a *≥*7 dB decline in average High-5 sensitivity during follow-up, compared to fewer than 30% when using MD, highlighting the superior sensitivity of High-5 to detect meaningful, localized deterioration. The 7 dB threshold itself is grounded in studies of perimetric test–retest variability, which demonstrated that glaucomatous progression could only be reliably distinguished from measurement noise when losses of this magnitude or greater occurred repeatedly.^24^ While such declines may take years to emerge in routine practice due to infrequent testing and measurement variability, time to detection can be substantially shortened with high-frequency or clustered testing. Combining these strategies with the High 5 approach may enable more timely identification of progression and facilitate the use of regulatorily aligned, clinically meaningful endpoints in future glaucoma trials.

An additional strength of the High-5 approach is its ability to enrich for eyes demonstrating rapid progression. In our analysis, nearly 90 percent of progressing eyes exhibited rates of change worse than 1 dB/year at High-5 locations, compared to substantially lower proportions when assessed using MD. Identifying such fast progressors is clinically meaningful, as these individuals are more likely to experience functional impairment and vision-related quality-of-life decline.^25,26^ While a rapid decline in MD reflects widespread visual field loss and may carry more immediate consequences for global visual function, localized deterioration at High-5 points can still have a profound impact, particularly when involving functionally critical regions, such as the central field.^27^ Also, such focal progression may signal aggressive disease that precedes broader functional decline. A binary endpoint based on the proportion of eyes exceeding a predefined rate threshold could also provide a patient-relevant and interpretable outcome to enhance trial feasibility.

It is important to emphasize that our model does not estimate absolute probabilities of whether an eye will progress. Given the retrospective nature of the data and variability in treatment decisions, such probabilities would be confounded by therapeutic interventions and disease management strategies. Prior attempts to predict who will progress using AI have yielded only modest accuracy.^28-30^ Instead, our model was designed to address the where question, i.e., identifying which locations within the visual field are most likely to deteriorate if progression were to occur. This spatial prioritization enables targeted monitoring and provides a data-driven framework for defining sensitive and clinically meaningful endpoints.

Our study has limitations. First, all performance estimates were derived from retrospective datasets. Although the external validation cohorts were independent, they were drawn from tertiary care centers, which may limit the generalizability of our findings to broader populations or clinical trial settings. Second, while the High-5 approach improves sensitivity by focusing on high-risk regions, its reliance on a limited number of visual field locations may introduce greater measurement variability. The choice of five points was guided by regulatory precedent, but because the model outputs relative probabilities for each test location across the entire visual field, it can be readily adapted to define endpoints based on alternative thresholds or numbers of locations (e.g., 7 or 10 points) tailored to specific trial designs or clinical scenarios. Finally, prospective studies will be important to assess the added value of integrating structural data, such as optical coherence tomography (OCT), into the model and to explore how the approach could be expanded or adapted to incorporate alternative testing paradigms, such as 10-2 visual fields.

In conclusion, we developed and validated an AI model capable of identifying visual field locations most likely to exhibit glaucomatous progression using baseline data alone. A derived High-5 metric, based on the five highest-risk locations per eye, demonstrated significantly faster rates of change and greater sensitivity for detecting progression compared to conventional metrics such as MD. These findings support the use of AI-informed, spatially targeted endpoints that align with regulatory priorities and offer a foundation for designing more efficient strategies to monitor glaucoma progression in both clinical practice and neuroprotection trials.

## Data Availability

The de-identified visual field data used in this study are available from the corresponding author upon reasonable request. Requests will be evaluated in accordance with institutional data-sharing policies.

